# Seroprevalence of Hospital Staff in Province with Zero COVID-19 Cases

**DOI:** 10.1101/2020.07.13.20151944

**Authors:** Tanawin Nopsopon, Krit Pongpirul, Korn Chotirosniramit, Wutichai Jakaew, Chuenkhwan Kaewwijit, Sawan Kanchana, Narin Hiransuthikul

## Abstract

**BACKGROUND:** COVID-19 seroprevalence data has been scarce, especially in less developed countries with a relatively low infection rate.

**METHODS:** A locally developed rapid immunoglobulin M (IgM) / immunoglobulin G (IgG) test kit was used for screening hospital staff in Ranong hospital which located in a province with zero COVID-19 prevalence in Thailand from April 17 to May 17, 2020. A total of 844 participants were tested; 82 of which were tested twice with one month apart. (Thai Clinical Trials Registry: TCTR20200426002)

**RESULTS:** Overall, 0.8% of the participants (7 of 844) had positive IgM, none had positive IgG. Female staff seemed to have higher IgM seropositive than male staff (1.0% vs. 0.5%). None of the participants with a history of travel to the high-risk area or a history of close contact with PCR-confirmed COVID-19 case had developed antibodies against SARS-CoV-2. Among 844 staff, 811 had no symptom and six of them developed IgM seropositive (0.7%) while 33 had minor symptoms and only one of them developed IgM seropositive (3.0%). No association between IgM antibody against SARS-CoV-2 status and gender, history of travel to a high-risk area, history of close contact with PCR-confirmed COVID-19 case, history of close contact with suspected COVID-19 case, presence of symptoms within 14 days, or previous PCR status was found. None of the hospital staff developed IgG against SARS-CoV-2.

**CONCLUSION:** COVID-19 antibody test could detect a substantial number of hospital staffs who could be potential silent spreaders in a province with zero COVID-19 cases. Antibody testing should be encouraged for mass screening, especially in asymptomatic healthcare workers.

**TRIAL REGISTRATION:** This study was approved by the Institutional Review Board of Chulalongkorn University (IRB No.236/63) and the Institutional Review Board of Ranong Hospital. (Thai Clinical Trials Registry: TCTR20200426002)

**FUNDING:** None.

## Introduction

Seroprevalence data has been scarce in Asian countries besides China. Along with the gold-standard polymerase chain reaction (PCR) testing, antibody testing is beneficial for epidemiological investigation and epidemic control of infectious diseases including the present coronavirus disease 2019 (COVID-19). In Singapore, serological evidence was used to trace and identify missing spreader for three clusters (1). Asymptomatic silent spreaders have been of major concern as suggested by a systematic review—could be as low as 1.6% of COVID-19 confirmed cases in China or as high as 51.7% of confirmed cases in Diamond Princess cruise (2).

Early COVID-19 prevalence studies were based only on PCR for the diagnosis of severe acute respiratory syndrome coronavirus 2 (SARS-CoV-2) infection in individuals. However, recent studies tended to report PCR along with serological test results. Our rapid systematic review of peer-reviewed evidence up to May 1, 2020, reported a range of seroprevalence among healthcare workers from 2.0% in Lebanon and Claremont, the USA to 4.5% in Padova, Italy (3). China reported an overall seroprevalence of 2.5% in a hospital setting, in which 1.8% and 3.5% were among healthcare workers and asymptomatic patients, respectively (4). Additionally, China studied the development of antibodies against SARS-CoV-2 in symptomatic confirmed COVID-19 cases and found that immunoglobulin M (IgM) reached its peak 20–22 days after onset while immunoglobulin G (IgG) reached its peak 17–19 days after onset (5). Some works emphasized on antibody testing for hospital workforce and policy issues. Another seroprevalence study in Belgium conducted on healthcare personnel who worked in a tertiary hospital found 6.4% seroprevalence and identified some risk factors for developing antibodies against SARS-CoV-2 (6).

More recent studies in the medRxiv preprint repository reported a range of seroprevalence in healthcare personnel from 0% (7) to 45.3% (8) with intra-regional variations. In China, for example, seroprevalence among hospital staff ranged from 1.8% (4) to 17.1% (9) whereas seroprevalence of healthcare workers in Asia besides China ranged from 3.3% (10) to 5.4% (11). In Southeast Asia, there was only a study from Thailand reported a 3.7% seroprevalence (12). Seroprevalences in healthcare personnel varied from 1.1% (13) to 45.3% in Europe (8) and from 2% (3) to 7.6% (14) in North America. Only one African study reported zero seroprevalences in hospital staff in Libya (7) whereas no seroprevalence studies were from South America, Antarctica, or Australia.

Ideally, both PCR and antibody testing provided complementing information to shape the picture of the situation in a specific hospital, area, or country. However, most low- to middle-income countries could not afford the cost of laboratory tests and had to develop criteria-based policies for resource-use optimization. In Thailand, for instance, the PCR is reserved for individuals who meet the national criteria for COVID-19 polymerase chain reaction testing.

Ranong is one of 77 provinces in Thailand with zero cumulative confirmed COVID-19 case from April 17 to May 17, 2020, and still had no case as of July 4, 2020 (Figure 1). Hospital is one of the highest risk areas for receiving and spreading pathogens—healthcare workers developed a higher chance of getting infected by co-workers or patients and vice versa. This study aims to estimate the hospital-wide seroprevalence in healthcare workers who worked in the largest public hospital in Ranong to develop the strategies to slow down the pandemic and ensure the safety of healthcare workers who come to work and patients who visit the hospital.

**Figure 1.**
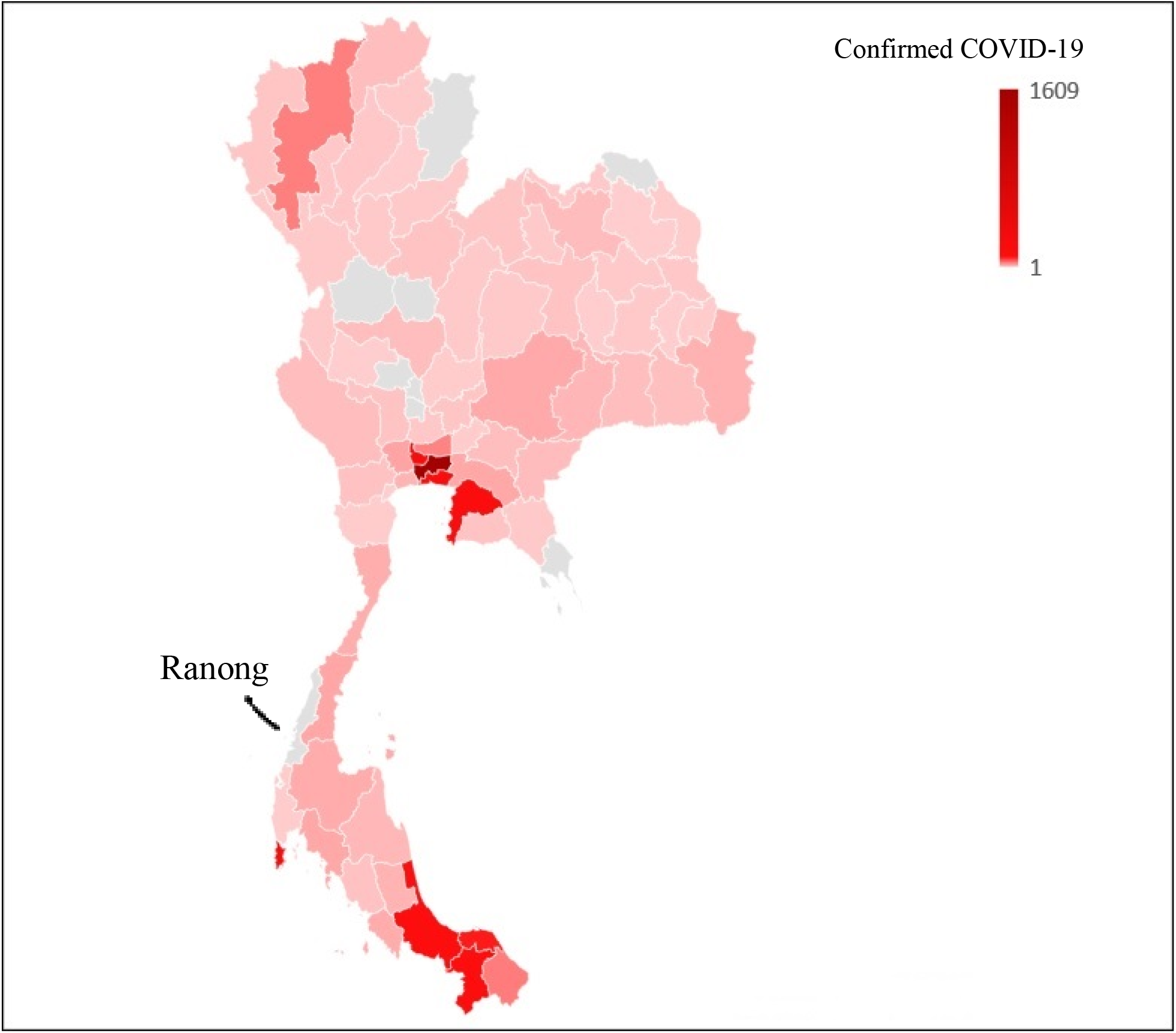
Numbers of Confirmed COVID-19 Cases across Geographical Areas of Thailand as of July 4, 2020. Bangkok Metropolitan had the most cumulative confirmed COVID-19 cases, 1,609 cases, while Ranong, the province of focus in this study, had no confirmed COVID-19 case.

## Results

### Healthcare workers’ demographics

All 844 Thai hospital staff were invited to participate in the study and tested for IgM and IgG antibodies against SARS-CoV-2 (100% participation rate). Their median age was 42 years (interquartile range 32–50). Most of them (71.7%) were female and 96.1% had no symptoms. The 33 symptomatic participants (3.9%) reported cough (1.7%), rhinitis (1.5%), sore throat (1.4%), dyspnea (0.5%), and fever (0.2%). History of travel to the high-risk area was 2.5%, history of close contact PCR confirmed COVID-19 case was 2.0%, history of close contact suspected case was 38.1%. Only 1% of participants had previous negative PCR results while the rest never get tested (Table 1).

### Serological results of healthcare workers in Ranong hospital

Overall, seven hospital staff tested positive for COVID-19 IgM (0.8%, 95% CI: 0.4%, 1.7%) while none of the participants developed IgG. Female staff seemed to have higher rate of positive IgM (1.0%, 95% CI: 0.5%, 2.1%) than male (0.5%, 95% CI: 0.1%, 2.6%). None of the participants with a history of travel to the high-risk area or a history of close contact with PCR-confirmed COVID-19 case developed antibodies against SARS-CoV-2. There was no statistical difference in IgM seroprevalence between staff with and without a history of close contact with suspected COVID-19 case. Among 844 staff, 811 had no symptom and six of them developed IgM seropositive (0.7%, 95% CI: 0.3%, 1.6%) while 33 had minor symptoms and only one of them developed immunoglobulin M (3.0%, 95% CI: 0.5%, 15.3%). Of 12 staff with sore throat, one had positive IgM (8.3%, 95%CI: 1.5%, 35.4%). There was zero IgM seroprevalence in staff with fever, cough, rhinitis, or dyspnea. Of 844 participants, eight had previous negative PCR result and none has developed the antibody for SARS-CoV-2 (Table 1). No association between IgM antibody against SARS-CoV-2 status and gender, history of travel to a high-risk area, history of close contact with PCR-confirmed COVID-19 case, history of close contact with suspected COVID-19 case, presence of symptoms within 14 days, or previous PCR status was found. None of the hospital staff developed IgG against SARS-CoV-2.

### Characteristics of seropositive participants

Seven participants developed IgM antibody in May. Their age ranged from 20 to 49 years. Six of them were female while only one staff was male. All of them were Thai, had no history of travel to a high-risk area, no history of contact with PCR-confirmed COVID-19 case, and no previous PCR status. Three had a history of contact with suspected COVID-19 case while the other three did not. Participant 7 was only one with IgM positive who had a symptom (i.e. sore throat) within 14 days before antibody testing. Nasopharyngeal swabs of all seven staff with a positive antibody tested negative for SARS-CoV-2 on PCR (Table 2).

### Repeating antibody tests in random sampling participants

Of 844 participants, 100 were randomly selected to get repeating antibody testing in April and May. None of them had any immunoglobulin developed in April whereas 82 also participated in antibody testing in May which showed no antibody against COVID-19.

## Discussion

In a small seaside province, Ranong, which had 193,370 inhabitants, located in the south of Thailand closed to the Andaman Sea and had border close to Myanmar, there was zero officially PCR-confirmed COVID-19 case compared to the current situation in Thailand (0.048 per thousand). This zero prevalence could be either from good compliance to public health recommendations or a low number of individuals who get PCR testing due to the stringent national criteria.

In this study, we reported a 0.8% IgM seroprevalence in Ranong hospital staff while their PCR tests were negative. While critics might argue that the antibody test had a high false-positive rate, especially in a population with high pre-test probability such as hospital staff, the PCR test could have a high false-negative rate, especially without a highly sensitive diagnostic test (15).

Of seven seropositive hospital staff, only one was symptomatic and none of them had a history of travel to high-risk area or history of close contact with PCR-confirmed COVID-19 case. The current national criteria and policies for PCR testing mostly rely on risk histories and symptoms so asymptomatic individuals or those with minor symptoms were left out, resulting in an underestimation of the actual prevalence of COVID-19.

Our study reported a lower seroprevalence in healthcare workers than hospitals in China (0.8% vs. 1.8%) (4), a tertiary hospital in Belgium (0.8% vs. 6.4%) (6), and a rural hospital located in low COVID-19 prevalence county of Germany (0.8% vs 2.7%) (16). Unlike China, Belgium, and Germany where the seroprevalences were mostly from positive IgG, our study revealed mostly positive IgM. Comparison with Belgium and Germany hospital should be interpreted with caution due to the unknown PCR status of subjects of the two studies.

COVID-19-confirmed patients developed peak IgM at 20–22 days and peak IgG at 17–19 days after symptom onset (5). For patients with no or minor symptoms, the onset might not be possible to determine. In this study, we used a qualitative antibody test kit so the relatively lower antibody level in early infected persons might not be detectable. Repeating PCR or quantitative antibody tests for suspected individuals was practiced in many countries. However, the cost would be doubled or more and might not be possible for low- or middle-income countries. We attempted to repeat the antibody test in randomly selected participants; however, only 82 of them could participate in the follow-up test and none of them developed antibodies against SARS-CoV-2 in both first and second tests. Rapid antibody tests should be more affordable to permit multiple tests among high-risk asymptomatic individuals who could be silent spreaders.

Serological testing provides some crucial epidemiological information and would have been more effective when combined with other diagnostic tests such as PCR. With immunoglobulin status and PCR results, we can shape the situation more precisely for both individual and regional views. Hopefully, with this and other vigorous and dedicated studies on antibody status around the globe, antibody testing would provide useful information for pandemic control.

In conclusion, the COVID-19 antibody test could detect a substantial number of hospital staffs who could be potential silent spreaders in areas with zero COVID-19 case. Antibody testing should be encouraged for mass screening, especially in asymptomatic healthcare workers.

## Methods

### Participants

All 844 healthcare workers including the physician, nurse, medical assistance, medical technician, and non-medical officers of the largest public hospital in Ranong province that reported zero cumulative PCR-confirmed COVID-19 cases during the entire study period (April 17 to May 17, 2020), were invited to participate in this study. All of them accepted to participate with written informed consent. Participants with active symptoms suiting national criteria for polymerase chain reaction testing were quarantined and excluded. Participants were asked to answer a survey about risk history for COVID-19, recent symptoms in the past two weeks, and previous PCR tests if available.

### Antibody testing

Locally developed Baiya Rapid IgG/IgM test kit (Baiya Phytopharm, Thailand) which reported the presence of IgM and IgG qualitatively, was used for antibody testing in this study. The internal validation of the test kit using the serum of 51 PCR confirmed COVID-19 cases and 150 controls showed sensitivity 94.1% (48 of 51) and specificity 98.0% (147 of 150) of IgM or IgG antibody. Participants with positive IgM were encouraged to have PCR test if available.

### Study procedures

On April 17, 2020, of 844 participants, 100 were randomly selected to have their first antibody testing. On May 17, 2020, all the remaining participants were tested for serological immunity whereas the 100 healthcare workers who tested in April had their second antibody testing.

### Statistics

Categorical data were presented with counts and percentages while continuous data were reported with median and interquartile range. Association between categorical variables and the status of immunoglobulin was analyzed using Fisher’s exact test. The 95% confidence interval (CI) of the seroprevalence was calculated by Wilson’s method using binomial probabilities. Missing data were excluded. All data were analyzed using Stata 16.1 (College Station, TX).

### Study approval

This study was approved by the Institutional Review Board of Chulalongkorn University (IRB No.236/63) and the Institutional Review Board of Ranong Hospital. (Thai Clinical Trials Registry: TCTR20200426002)

## Data Availability

Data is available from the corresponding author upon request.

## Author Contributions

TN, KP, and NH conceptualized the study, designed methodology, and administrated the project. TN, KC, KP, WJ, CK and SK contributed to data curation and clinical investigation. TN, KC, and KP validated the data. TN and KP performed formal analysis and provided software. TN visualized the data. KP, WJ, SK, NH provided resources for the study. KP, SK, and NH supervised the study. TN and KP wrote the original draft of the manuscript. All authors reviewed and edited the final manuscript.

## Acknowledgements

We thank Baiya Phytopharm, Thailand for supporting the Baiya Rapid COVID-19 IgG/IgM test kit. The company did not involve in the data analysis, interpretation, or manuscript preparation.

